# The Current Understanding of Precision Medicine and Personalised Medicine in Selected Research Disciplines – Study Protocol of a Systematic Concept Analysis

**DOI:** 10.1101/2022.01.12.22269125

**Authors:** Nicola Brew-Sam, Anne Parkinson, Christian Lueck, Ellen Brown, Karen Brown, Anne Bruestle, Katrina Chisholm, Simone Collins, Matthew Cook, Eleni Daskalaki, Janet Drew, Harry Ebbeck, Mark Elisha, Vanessa Fanning, Adam Henschke, Jessica Herron, Emma Matthews, Krishnan Murugappan, Dragomir Neshev, Christopher Nolan, Lachlan Pedley, Christine Phillips, Hanna Suominen, Antonio Tricoli, Kristine Wright, Jane Desborough

## Abstract

**Introduction:** The terms “precision medicine” and “personalised medicine” have become key terms in health-related research, and in science-related public communication. However, the application of these two concepts and their interpretation in various disciplines are heterogeneous, which also affects research translation and public awareness. This leads to confusion regarding the use and distinction of the two concepts.

**Methods and analysis:** Our study aims at using Rodger’s concept analysis method to systematically examine and distinguish the current understanding of the concepts “precision medicine” and “personalised medicine” in clinical medicine, biomedicine (incorporating genomics and bioinformatics), health services research; physics, chemistry, engineering; machine learning, and artificial intelligence, and to identify their respective attributes (clusters of characteristics) and surrogate and related terms. We will analyse similarities and differences in definitions in the respective disciplines and across different (sub)disciplines. The analysis procedure will include (1) a concept identification, (2) a setting, sample, and data source selection, (3) data collection, (4) data analysis and data summary, (5) identification of examples, and (6) identification of implications for further concept development.

**Ethics and dissemination:** Following ethical and research standards, we will comprehensively report the methodology for a systematic analysis following Roger’s[1] concept analysis method. Our systematic concept analysis will contribute to the clarification of the two concepts and distinction in their application in given settings and circumstances. Such a broader concept analysis will contribute to non-systematic syntheses of the concepts, or occasional systematic reviews on one of the concepts that have been published in specific disciplines, in order to facilitate interdisciplinary communication, translational medical research, and implementation science.

**Strengths and limitations of this study:**

- In contrast to previous studies, we examine the definitions of “precision medicine” and “personalised medicine” in specific selected disciplines in order to facilitate interdisciplinary communication, translational medical research, and implementation science.
- Moreover, we analyse these two concepts systematically and base our review on renowned concept analysis methodology.
- Our study contributes to the clarification of the two concepts, their attributes, and differences in various disciplines.
- Concepts are constantly developing and their meanings change over time, and hence it is not possible for our analysis to deliver an unequivocal definition.

## Introduction

The terms “precision medicine” and “personalised medicine” are increasingly used in health-related research. However, the interpretation of these concepts and their application in various disciplines are heterogeneous[2, 3]. The terms are often used in relation to specific diseases (e.g., cancer) with an applied focus but without a detailed definition or specification of the underlying concepts. Non-systematic syntheses of (one of) the concepts[4, 5], or occasional systematic reviews on (one or other of) the concepts have been published in specific disciplines[6, 7]. But, so far, understanding of the concepts has not been systematically clarified and compared across disciplines. Erikainen and Chan[4] report that disciplines have inconsistent, and potentially contrary, understandings of precision medicine or personalised medicine, as well as differing preferences of terms describing similar entities (for an exemplary variety of definitions see **Table 1**).

**Table 1.**
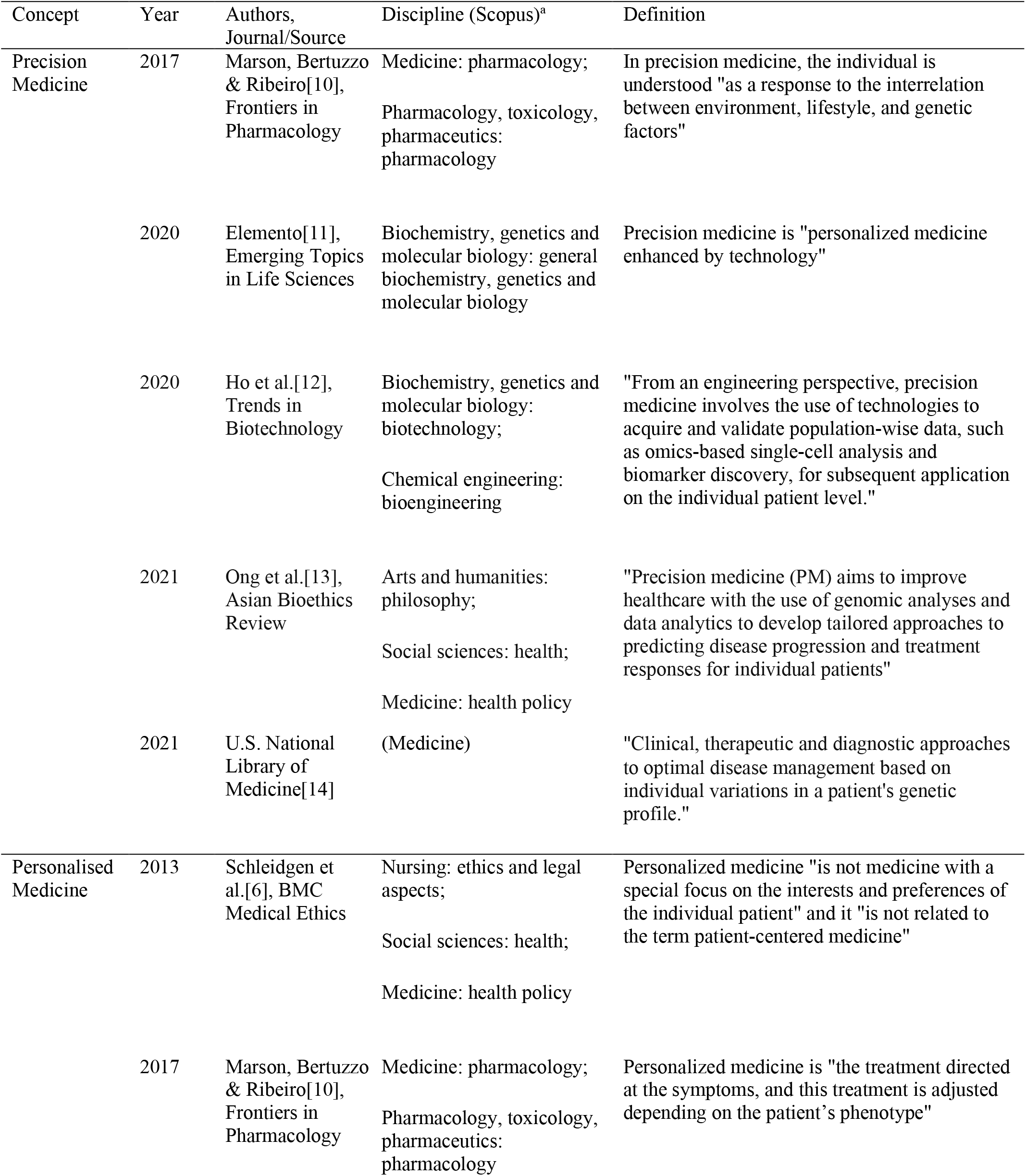
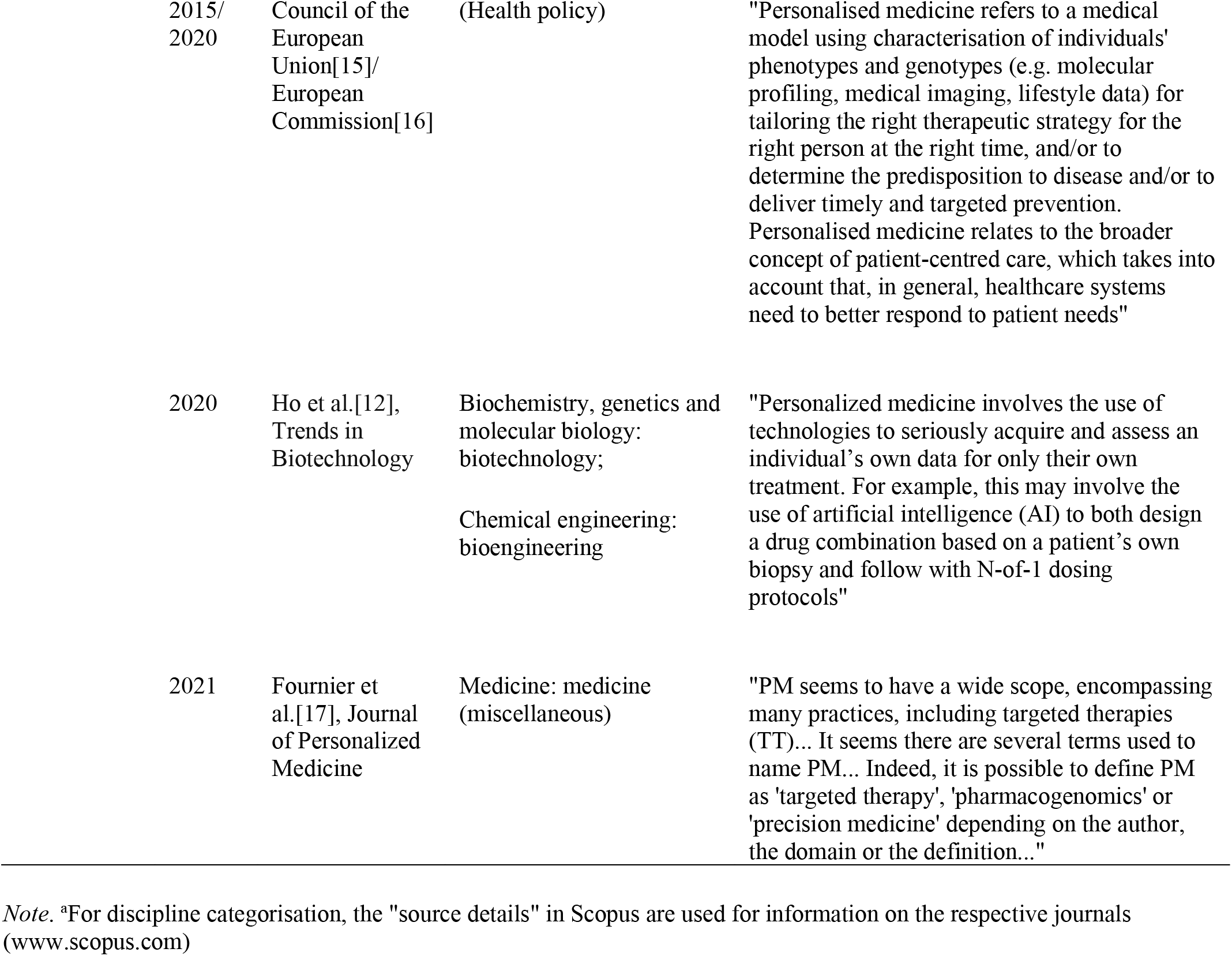
Variety of definitions of “precision medicine” and “personalised medicine” – examples

For example, discussion in biomedical literature has relatively early started to express criticism of the term “personalised medicine” arguing that this term implies unrealistic optimism and promises in relation to what biomedical technology will be able to deliver. According to Schleidgen et al.[6] “personalised medicine” wrongly implies a focus on the interests and preferences of an individual and the provision of patient-centered medicine through personalised medicine alone. A similar statement was put forward by the United States National Academies[8]. Schleidgen et al.[6] recommended the use of the term “stratified medicine” instead of “personalised medicine” to present a more realistic vision of the underlying concept[4]. Similarly, “precision” promises a level of certainty[9] “that is unlikely to be reflected in the realities of precision medicine”[8].

However, other disciplines (including medical ethics, critical studies of biosciences, patient-centered research) have criticised Schleidgen’s view, suggesting it can only be applied in a biomedical context, emphasizing that “personalised medicine” should be clearly distinguished from “patient-centered care”[4]:

> Schleidgen and colleagues’ definition can be seen as privileging the biological and molecular interpretation of “personalization” as group-level treatment stratification, to the explicit and purposeful exclusion of patient-centered interpretations.[4]

To summarise, the understanding and use of the concepts “personalised” and “precision” medicine is discipline-dependent, which may also influence discussions on a policy level[15, 18-20]. The latter can be shown by country-specific preferences at a policy level. For example, while the discussion is mostly focusing on precision medicine in the U.S. since the Precision Medicine Initiative was launched in 2015[19], the European Union set up an International Consortium for Personalised Medicine (ICPerMed) in 2016[16]. Yet, the distinction of the two concepts is not clear enough to argue how the two terms differ. Preferences and an understanding largely depend on historical developments of the terms in specific contexts, leading to varying interpretations (e.g., on a research and a policy level). This variety in interpretations, going along with the use of different terms, can have negative consequences, including the creation of different representations or beliefs in people when using a variety of terms, or a reinforcement of inaccuracies in definitions[17].

A first step towards more clarity is an understanding of current differences and similarities regarding precision and personalised medicine across disciplines in research. Further research can then examine how these current interpretations developed historically (concept revisions), and how they influence(d) policy.

Our Health in Our Hands (OHIOH) is a multidisciplinary research project at the Australian National University which brings together the disciplines of clinical medicine, biomedicine (incorporating genomics and bioinformatics), health services research; physics, chemistry, engineering; machine learning, and artificial intelligence. These disciplines are the focus of this paper. OHIOH aims to advance digitalisation and personalisation of healthcare using a co-production approach with partners from research, lived experience, healthcare professionals, and health services[21]. OHIOH research includes studies on the experiences of people living with Multiple Sclerosis (MS)[22] and Type 1 Diabetes[23]. A recently published OHIOH paper[24] discusses the understanding and experiences of people living with MS in order to emphasise the importance of personalised medicine in MS treatment and care. In the course of drafting the manuscript, discussions around “personalised” and “precision” medicine revealed that the understanding of these two concepts varied greatly between the researchers in the various disciplines involved in OHIOH.

### Objective

The aim of this study is to examine the current understanding of the concepts “precision medicine” and “personalised medicine” in clinical medicine, biomedicine (incorporating genomics and bioinformatics), health services research; physics, chemistry, engineering; machine learning, and artificial intelligence using Rodgers’[1] concept analysis to identify concept attributes (clusters of characteristics) and to determine how these two concepts are distinguished in these selected disciplines and potential subdisciplines. We will answer the following research questions (RQ):

RQ1: What is the current understanding of “precision medicine” and “personalised medicine” in clinical medicine, biomedicine (incorporating genomics and bioinformatics), health services research; physics, chemistry, engineering; machine learning, and artificial intelligence, and what are similarities and differences in definitions in the respective disciplines and across different (sub)disciplines?

RQ2: What are the related and surrogate terms for “precision medicine” and “personalised medicine” in each of the selected disciplines?

It is important to understand how the two concepts are currently interpreted and understood in the individual disciplines in order to be able to create consistent understanding of the concept interpretations and to reduce ambiguity in the literature. This current interpretation does not include concept revisions over time which could be examined in follow-up research.

## Methods and Analysis

### Protocol development

We used the 17 Preferred Reporting Items for Systematic Review and Meta-Analysis Protocols (PRISMA-P) throughout the development of our study protocol[25] (see supplementary file). Our systematic data collection and analysis will similarly follow the subsequent Preferred Reporting Items for Systematic Reviews and Meta-Analyses (PRISMA)[26]. Amendments to the study protocol will be reported in the final published systematic concept analysis manuscript.

### Concept analysis approach

A concept analysis aims to clarify a concept (e.g., attributes, antecedents, consequences)[1]. Such a concept clarification enables assessment of a concept’s strengths and weaknesses[27]. Based on Walker and Avant’s[28] traditional approach^1^ derived from Wilson’s method[29] which is based on realism (deductive analysis), Rodgers[1] developed an evolutionary concept analysis based on relativism (inductive analysis). Rodgers[1] viewed concepts as dynamic and evolving phenomena without strict boundaries. This took account of the fact that concepts are constantly developing and their meanings change over time, and hence it is not possible for an analysis to deliver an unequivocal definition[27]. Moreover, concepts are understood differently in different disciplines due to what Rodgers[1] calls “enculturation” within individual disciplines. Thus, it is important to clarify the selection of disciplines being focused on when using Roger’s approach to a concept analysis.

Bearing in mind the changing and non-static understanding of the terms “precision medicine” and “personalised medicine” over time, Rodger’s approach appears to be a good fit for analysis. Our analysis of “personalised” and “precision” medicine will only represent a snapshot of the current understanding of these concepts in the literature pertaining to clinical medicine, biomedicine (incorporating genomics and bioinformatics), health services research; physics, chemistry, engineering; machine learning, and artificial intelligence. Nevertheless, it is essential to highlight any differences and similarities between disciplines to inform current and future research, aiming to standardise and generate a uniform approach if at all possible. We will not look into revisions of definitions over time, the focus is on the current understanding of the two concepts.

Rodger’s[1] concept analysis has generated a lot of attention in the healthcare context, and has become a recognised method of concept clarification. For example, Hudon et al.[30] used Rodger’s concept analysis approach to analyse “enablement” in a healthcare context. In Rodger’s method of concept analysis[1], concepts are considered to be abstractions that are expressed in an arbitrary form. They constitute a mental (re)grouping of a number of attributes. Hudon et al.[30] define attributes as characteristics of concepts that must be present for the recognition of the concept as an entity. Concept analyses are employed in developing valid measuring instruments which can evaluate the attributes of a concept (determining whether there is good content validity)[30].

### Concept analysis procedure

Rodger’s[1] concept analysis is divided into six steps, comprising (1) the identification of the concepts of interest and associated expressions and background, (2) the selection of an appropriate realm for data collection (setting, sample and data sources), (3) the collection of data relevant to identifying concept attributes and the contextual concept basis, (4) the analysis and data summary regarding the concept characteristics, (5) the identification of concept examples, and (6) the identification of implications for further concept development.

#### Concept identification (step 1)

By way of example, Viana et al.[31] state that “precision medicine” and “precision health” are not identical:

> Distinct from precision medicine, precision health takes a lifespan perspective in health monitoring, identifying actionable risks and intervening early.[31]

Our systematic concept analysis will focus on the clarification of the two concepts “precision medicine” and “personalised medicine”. Surrogate and related terms will not be identified in advance: surrogate terms are other terms used to describe identical concepts, while related terms describe entities that are not identical but have something in common with the concepts under analysis[1]. The exploration of these will be conducted at a later step in the full text analysis of the included papers[compare 32]. Related (but not identical) concepts such as individualised care, stratified medicine, P4 medicine (predictive, preventive, personalised and participatory), genomic medicine, or patient-centered care, will be collected in the systematic review alongside definitions and interpretations derived from analysis of the full texts. Similarly, replacement terms for “medicine” in “precision/personalised medicine” such as “health(care)”, “treatment”, “therapy/therapeutics”, “medical care”, or similar composite terms[33] will be collected during full text analysis. Since related concepts are not identical with precision or personalised medicine, these are not central to the focus of this study. The main focus will be on the two terms “precision medicine” and “personalised medicine”.

#### Setting, sample, and data source selection and data collection (steps 2 and 3)

As above, the disciplines of clinical medicine, biomedicine (incorporating genomics and bioinformatics), health services research; physics, chemistry, engineering; machine learning, and artificial intelligence were selected for analysis due to their key roles in OHIOH, which is an interdisciplinary research collaboration. The analysis might also reveal diverse understandings of the concepts in subdisciplines of the selected disciplines. This will be considered adequately in the analysis.

The search strategy will be used to search a number of databases relevant to each of these disciplines. We will use the databases PubMed/Medline, CINAHL, Scopus, Web of Science, Google Scholar, Research Square, F1000 Research and other computing-specific databases listed in **Table 2**. These are among the most representative and commonly used databases for the included disciplines.

**Table 2.** Collection of databases for the selected disciplines

| Database | Disciplines |
| --- | --- |
| CINAHL | Clinical medicine, health services research |
| F1000 Research | Biomedicine |
| Google Scholar | Clinical medicine, health services research, physics, chemistry, engineering, machine learning and artificial intelligence |
| https://aclanthology.org/ | Machine learning and artificial intelligence |
| https://arxiv.org/ | Machine learning and artificial intelligence |
| https://dblp.org/ | Machine learning and artificial intelligence |
| https://dl.acm.org/ | Machine learning and artificial intelligence |
| https://paperswithcode.com/ | Machine learning and artificial intelligence |
| https://www.ieee.org/ | Machine learning and artificial intelligence |
| PubMed/Medline | Clinical medicine, biomedicine, health services research, medical informatics |
| Research Square | Biomedicine |
| Scopus | Clinical medicine, health services research, medical informatics, physics, chemistry, engineering |
| Web of Science | Clinical medicine, health services research, physics, chemistry, engineering |

Once identified, the relevant discipline of a given publication will be defined according to the chosen publication’s profile in Scopus (www.scopus.com). Scopus delivers a detailed categorisation and classification of journals into disciplines.

Additional manual hand searching will be carried out to identify potentially relevant articles that might have been missed in the searches of the above databases (e.g., references of papers).

The search strategy will look for articles that mention “precision medicine” and/or “personalised medicine” in their titles in addition to “defin*” (definition/define) or “concept*” in the full text. Both British and American spelling will be accepted (“personalised”/ “personalized” medicine). As an example, a PubMed search string for precision and personalised medicine will include:

> ((precision medicine[Title]) OR (personalised medicine[Title]) OR (personalized medicine[Title])) AND ((defin* [Text Word]) OR (concept* [Text Word]))

The search will be limited to scientific research papers in English language published in peer-reviewed academic journals. Moreover, the search will be limited to articles published from 2016 to 2022 in order to capture the current understanding of these concepts following the introduction of major initiatives such as the Precision Medicine Initiative[19] or the International Consortium for Personalised Medicine in 2015/2016[16].

Any articles with a main focus on clarifying at least one of the concepts and contributing to a deeper understanding of the concept(s) will be included (**Table 3**). Articles that do not offer any substantial (theoretical) basis underlying the clarification of the concepts will be excluded. Empirical studies will be included if they serve the purpose of concept clarification (e.g., hybrid concept analysis which combines empirical research/fieldwork such as expert interviews with the analysis of a concept[34]).

**Table 3.** Eligibility criteria for articles

|  |
| --- |
| • Publications in peer-reviewed academic journals |
| • Published in the disciplines of clinical medicine, biomedicine (incorporating genomics and bioinformatics), health services research; physics, chemistry, engineering; machine learning, and artificial intelligence (discipline defined by the chosen publication's profile in Scopus) |
| • Published between 2016 and 2022 (including papers under review) |
| • Published in English language |
| • Having a main focus on clarifying at least one of the concepts "precision medicine" or "personalised medicine", and contributing to a deeper understanding of the concept(s) using theoretical or empirical studies – publications that do not deliver any substantial contribution regarding the clarification of the concepts are to be excluded |
| • Empirical studies will be included if they serve the purpose of concept clarification (e.g., hybrid concept analysis which combines empirical research with the analysis of a concept) |

The study selection procedure will follow standard practice, i.e. an initial first search by one researcher who will remove any duplicates, followed by screening of titles and abstracts, and then full article screening, by several researchers from the included disciplines. Any disagreements regarding article inclusion will be resolved through discussion with additional researchers.

Search and inclusion results will be displayed using a PRISMA flow diagram. A search/study log book will be used to support study reliability estimation, with notes about the search and data collection procedure taken by the researchers throughout the data collection.

Data will be managed using a reference manager (Endnote), an Excel list with study details and data extraction summaries, and the systematic review management program Covidence to organise data collection and analysis steps.

#### Analysis, data summary, and identification of examples (steps 4 and 5)

For full text analysis, every included article will be read with a focus on extracting information relevant to the interpretation and definition of the two concepts, as well as their contextual basis, their attributes and any related and/or surrogate terms (**Table 4**). Data extracted from the eligible papers will include the journal name, research discipline/context (including subdiscipline), authors, year, citation, study aim, and definitions of the concepts, attributes/characteristics, concept differences/similarities, related and surrogate terms, quality appraisal, and further notes. For empirical studies, risk of bias will be assessed using the Mixed Methods Appraisal Tool (MMAT)[35].

**Table 4.** Questions to explain the categories for analysis

|  |  |
| --- | --- |
| Concept attribute | What are the concept's characteristics? |
| Concept definition | Which definitions are presented in the data material? |
| Concept differences/similarities | Are specific concept differences or similarities mentioned in the data material? |
| Contextual basis | In which context is the concept presented (research discipline)? |
| Example | Are examples of the concept described in the data material? |
| Related term | Do other words have something in common with the concept? |
| Surrogate term | Do other words say the same thing as the chosen concept? |

*Note.* Based on Tofthagen and Fagerstrom[27]

Several researchers with expertise in cooperating across the included disciplines (who participated in the abstract and full article screening) will initially extract data from 5-10 papers for the extraction to be compared in order to reach consensus for the further extraction. The subjectivity of the views of involved researchers will be identified and discussed through inter-coder comparisons, and differences will be resolved through discussion with additional researchers, if necessary. In the following, data will be extracted from all remaining included articles by the same researchers.

Rodgers’ concept analysis method will guide a narrative synthesis, an example can be found in Miles and Huberman[36]. The findings will be compared, with (dis)similarities within the disciplines analysed[27]. Through this process, patterns will be revealed and main themes will be identified. This is a continuous process of data analysis, the data being reorganised until a descriptive pattern of themes is reached[1]. A data summary requires consensus regarding the concepts and their attributes, and (practical) examples from the included articles are used for concept explanation and illustration[1, 27]. Several researchers will be involved in the data analysis in order to avoid subjective interpretation.

#### Identification of implications for further concept development (step 6)

Rodgers[1] suggests that, based on the data analysis, further questions and hypotheses are presented, rather than an attempt to generate a ‘final’ definition of the concepts involved, since this is not possible due to the dynamic nature of concepts. Suggestions will be made in relation to directing future research and helping guide further concept development and analysis.

### Patient and public involvement

OHIOH is a project that spans several disciplines and is underpinned by a participatory co-production model of research that is characterised by close collaboration with clinicians and with people living with Multiple Sclerosis or Type 1 Diabetes in research planning and in conducting the research. Our co-production partners have been involved in the development of this protocol and will continue to be involved throughout the research project. After finalising the data collection and data analysis, the results will be discussed with our partners and the OHIOH teams in order to put the results in an OHIOH context. For each discipline, the respective OHIOH team will review the resulting understanding of personalised and precision medicine from their scientific perspective.

### Ethics and dissemination

Following ethical and research standards, we will comprehensively report the methodology for a concept analysis following Rodgers[1]. Ethical approval is not required for this research because our study collects publicly available and theoretical data about the concepts underlying “precision medicine” and “personalised medicine”. The results of this study will be disseminated through publication in peer-reviewed academic journals and at scientific conferences. Our findings will contribute to clarification of the underlying concepts and so help guide future research.

## Potential study limitations

Potentially relevant articles might be missed using any specific search strategy. However the inclusion of several databases and additional hand searching as part of a systematic approach is likely to minimise the risk of missing significant literature. Difficulties arising in an application of concept analysis to precision/personalised medicine could include that in contrast to other theoretical concepts (e.g., enablement), precision/personalised medicine are mostly practical medical terms rather than mere theoretical concepts.

## Data Availability

All data that will be produced in the present study will be available upon reasonable request to the authors.

## Acknowledgements and funding

This research was funded by and has been delivered in partnership with Our Health in Our Hands (OHIOH), a strategic initiative of the Australian National University, which aims to transform health care by developing new personalized health technologies and solutions in collaboration with patients, clinicians, and health care providers. AT gratefully acknowledges the support of the Australian Research Council (ARC) (DP190101864 and FT200100939) and NATO Science for Peace and Security Program AMOXES.

## Authors’ Contributions

NBS prepared the study protocol and drafted the manuscript. JD supervised the procedure of developing the study protocol. All authors were involved in study concept and design discussions. All authors were involved in manuscript revision.

## Footnotes

1 Walker & Avant (2019) view concepts as static entities which are independent of context and have clear boundaries.

